# Incidence of Microbial Infections in English UK Biobank Participants: Comparison with the General Population

**DOI:** 10.1101/2020.03.18.20038281

**Authors:** Bridget Hilton, Daniel Wilson, Anne-Marie O’Connell, Dean Ironmonger, Justine K Rudkin, Naomi Allen, Isabel Oliver, David Wyllie

## Abstract

Understanding the genetic and environmental risk factors for serious bacterial infections in ageing populations remains incomplete. Utilising the UK Biobank (UKB), a prospective cohort study of 500,000 adults aged 40-69 years at recruitment (2006-2010), could help address this.

We assess the feasibility of linking an England-wide dataset of microbiological isolations to UKB participants, to enable characterisation of microbial infections within the UKB Cohort. Microbiological infections occurring in patients in England, as recorded in the Public Health England Second Generation Surveillance System (SGSS), were linked to UKB participants using pseudonymised identifiers. By January 2015, ascertainment of laboratory reports from UKB participants by SGSS was estimated at 98%. 4.5% of English UKB participants had a positive microbiological isolate in 2015. Half of UKB isolates came from 12 laboratories, and 70% from 21 laboratories. Incidence rate ratios for microbial isolation, which is indicative of serious infection, from the UKB cohort relative to the comparably aged general population ranged from 0.6 to 1, compatible with the previously described healthy participant bias in UKB.

Data on microbial isolations can be linked to UKB participants from January 2015 onwards. This linked data would offer new opportunities for research into infectious disease in older individuals.

## Introduction

### Infection incidence rises in older individuals

Bacterial infection is an important cause of death in older individuals ^1^. Infection can be diagnosed based on clinical presentation, although symptoms are less predictive of bacterial infections in older individuals ^1^. The addition of radiological imaging (e.g. chest X-rays) can provide indication of infection ^2,3^, but radiological diagnosis is unusual in primary care, at least in the UK. Microbiological sampling can also contribute to the diagnosis of infection, and reveal the causative organism: a diagnosis of severe bacterial infection can be made following microbiological culture of normally sterile sites, including blood, peritoneal fluid, and cerebrospinal fluid ^4,5^. Positive microbiological cultures of urine are also commonly associated with infection ^6,7^.

Incidence rates of bacterial infections increase markedly with age. For example, English surveillance data shows that the incidence of *E. coli* bacteraemia is more than ten-fold higher in 45-64 year old men, and about 100 fold higher in over 75 year olds ^8^, compared with 15-44 year olds. Similar trends are observed with *S. aureus* ^9^, *S. pyogenes* ^10^ and *S. pneumoniae* ^10,11^ bacteraemia. The age-associated increased incidence of severe infection is observed both in individuals with healthcare exposure and in individuals without prior hospital exposure ^12^. Marked age-associated increases in infection rates are also observed in community-origin conditions diagnosed syndromically in general practice, such as respiratory infections^13^.

### The determinants of age-related rise in infection incidence

The reasons for the age-related increase in infection incidence are not fully understood. Possible contributing factors include environmental risk factors, including housing, nutrition and other aspects of lifestyle. There is well documented population variability in innate immune function, e.g. in baseline inflammatory activity as reflected in serum CRP concentrations^14^, and changes in macrophage function associated with vitamin D metabolites^15^ which may be relevant to individual infection risk. Age-specific declines in adaptive immunity (e.g. T cell responses, antibody concentrations) may also be relevant, and have been shown, using sero-epidemiological and vaccination studies, to be related to the age-related increase in pneumococcal pneumonia and herpes zoster infection ^16,17^. Finally, germline genetic polymorphisms that predispose to infection^18-20^ may be revealed, as environmental predispositions increase.

### Assessing environmental, immune and genetic contributions of infection incidence

Assessing the impact of environmental, innate and adaptive immune, and genetic risk factors for infection in older adults is complex. For the comprehensive and reliable quantification of the combined effects of lifestyle, environment, genes and other exposures on a range of infectious diseases, prospective studies have a number of important advantages over retrospective case-control studies:

- They allow a wide range of different infectious diseases to be studied.
- Exposures can be assessed prior to disease development, which usually improves detail and accuracy (reduced variance) of information. Causal interpretations of associations between prior exposures and subsequent outcomes may be more robust (or enriched for true positives) because anachronous associations are not hypothesised, compared to other retrospective case-control studies.
- Investigation becomes possible into disease risk factors that might be affected by infection and/or its treatments (e.g. immune status blood marker concentrations, gut microbiome/metabolomics) or by an individual’s response to developing a bacterial infection (e.g. weight, physical activity, diet).
- Prospective studies are also able to better assess severe infections that have a high case-fatality rate, as such cases cannot readily be studied retrospectively.
- Prospective studies enable research into how infectious disease ‘exposure’ is related to a wide range of subsequent chronic health conditions.

However, because the incidence of severe infection in the general population is low (*E. coli* blood stream infection, which is the commonest blood stream infection in the UK, has an incidence of about 50 per 100,000 in 45-64 year olds), then prospective studies investigating infection risk factors in the general population need to be large.

### The UK Biobank

The UK Biobank (UKB) study is a prospective cohort study, which recruited around 500,000 men and women aged 40-69 years who lived within travelling distance of one of 22 recruitment centres between 2006 and 2010 ^21^. It was designed to assess the genetic and environmental determinants that contribute to common life-threatening and disabling diseases ^21^. Of the 500,000 participants recruited to UK Biobank, 445,023 (89%) were resident in England, heterogeneous and focused on certain major population areas (Figure 1).

**Figure 1:**
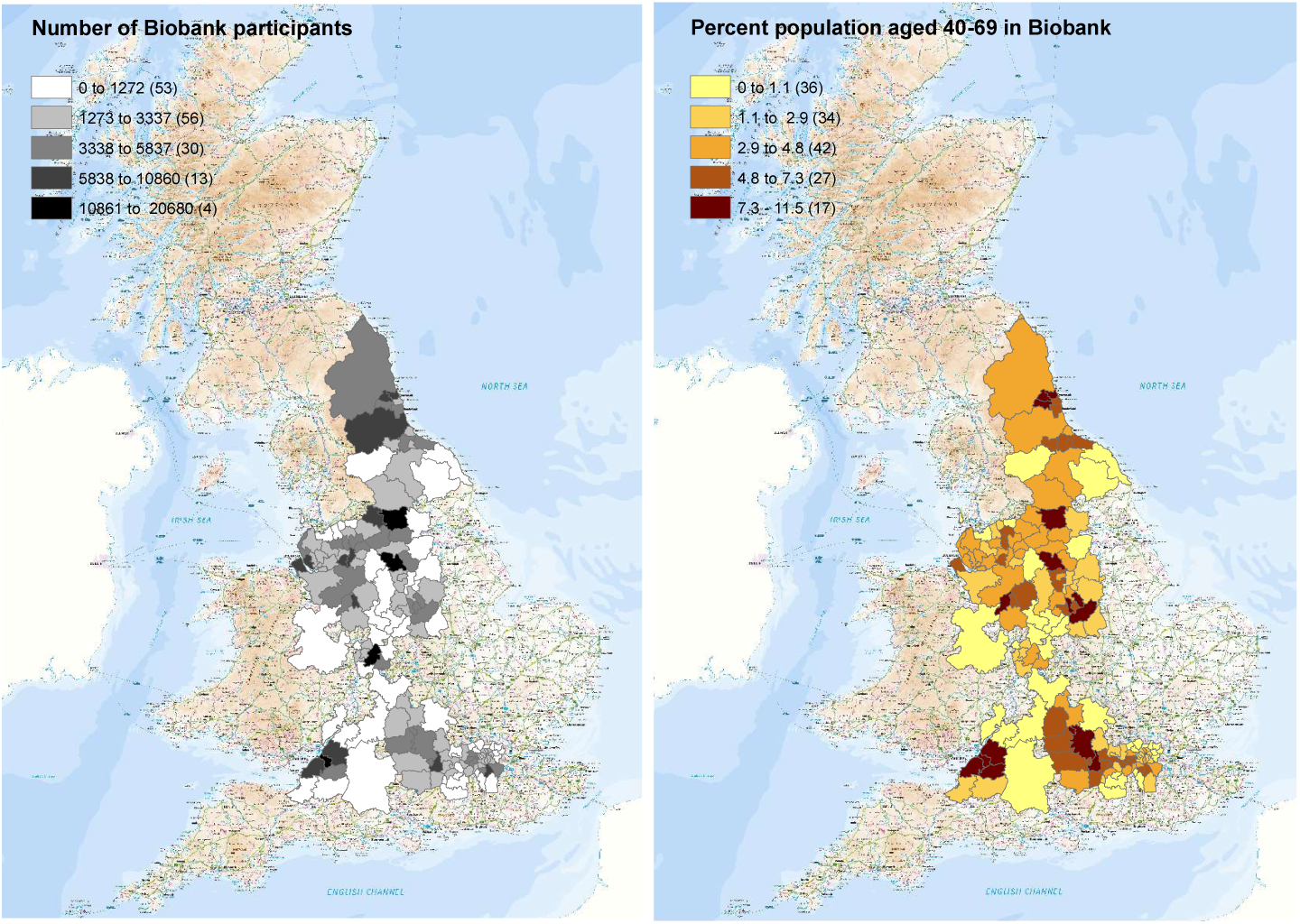
The numbers of Biobank participants identified in each local authority in which recruitment occurred (left), and the percentage of the population in each local authority recruited (right).

In addition to providing baseline health data and biological samples for biomarker measurement and genotyping, participants continue to be invited to undertake ongoing enhancements (e.g., multi-modal imaging, physical activity monitoring, completion of a series of web-based questionnaires). All participants also consented to linkage of their health records, such as death, cancer, hospital inpatient records and primary care data ^22^. Therefore, the UK Biobank cohort is one potential setting in which study of the determinants of microbial infection and of the sequelae of infection (including death) could be carried out, if it is possible to link UK Biobank participants to laboratory records of microbial isolation.

### The Second Generation Surveillance System (SGSS)

In England, the processing of microbiological specimens predominantly occurs in hospital laboratories run by the National Health Service. As part of its role in monitoring and improving population health, Public Health England has established a database (SGSS) that includes details of all positive microbiological isolates (microbial cultures) on which antimicrobial susceptibility testing is performed in all NHS Trusts in England. Near universal coverage was achieved by 2015, following work undertaken in support of the UK Government’s Antimicrobial Resistance Strategy ^22^. The database is a cornerstone of PHE surveillance, with aggregated information on bacterium/resistance profile combinations (‘bug-drug combinations’) ^22^ being fed back to the healthcare system via a web based portal in an effort to tailor prescribing to local resistance patterns ^7^

Here, we report the results of a pilot project linking data from UKB to the microbial isolations reported in SGSS and provide a description of the patterns of bacterial infection in the UKB cohort compared with the general population.

## RESULTS

### NHS numbers are sufficient for data linkage of microbiology samples in England

Between 1 April 2010 and 30 June 2016, data on microbial isolates from 4,726,417 samples (in 4,066,974 individuals aged 40-69 years) were deposited within SGSS (Figure 2A). The majority of samples had NHS number (exceeding 90% across all years of analysis, but increasing to over 98% from 2015 onwards, because using NHS numbers in laboratory test requests was mandated by statute in 2014). A change in the nature of the data feeds into SGSS during 2014 also led to a marked increase in availability of surnames from 2015 onwards (Figure 2B).

**Figure 2:**
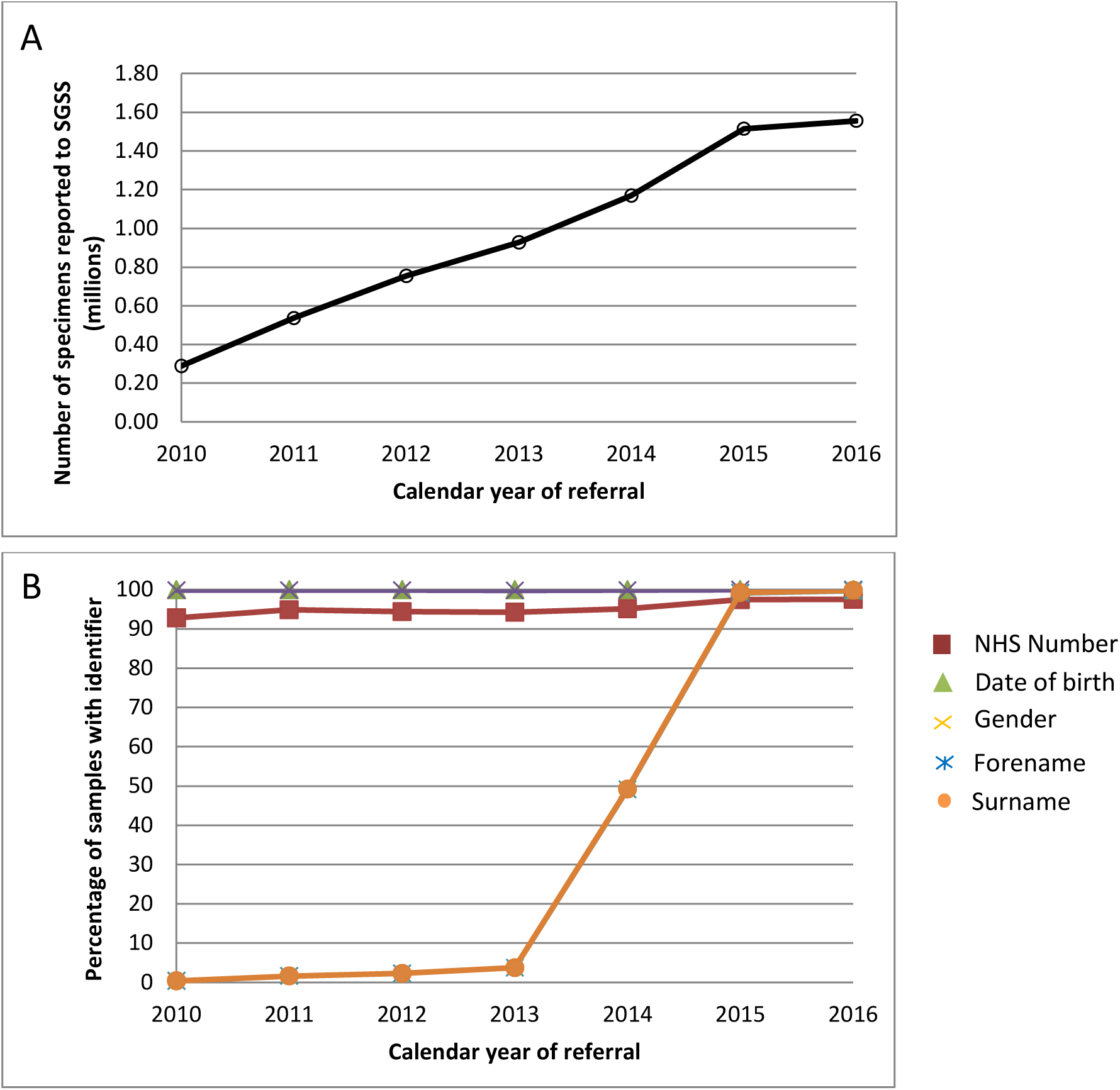
The numbers of specimens reported to SGSS (A) and the percentage of those specimens with various identifiers (B).

We considered whether linkage using additional identifiers (surname, forename, date birth) increased the proportion of linked records, and how this affected the specificity of linkage, i.e. the proportion of records falsely linked to an individual. We computed mean number of unique NHS numbers for sets of records putatively belonging to an individual, as identified by various composite identifiers made up of forename, surname, and date of birth. These investigations indicated that use of composite identifiers (in addition to NHS numbers) were likely to link records from individuals other than the intended individual. Given the almost universal nature of NHS number use post 2015, and the statutory requirement that the NHS uses it going forward, we conducted subsequent record linkage using NHS number alone.

### Near complete coverage of English UKB microbiological isolation by 2015

Numbers of samples arriving in SGSS increased year-on-year 2010 to 2015, before stabilising (Figure 2A). This increase coincided with a PHE initiative to encourage and assist all laboratories in England to report routinely to SGSS; by 2015, only three out of 172 laboratories were not doing so.

Based on the proportion of laboratories reporting to SGSS, the local authority areas in England they cover, and the residence of UK Biobank participants, the estimated ascertainment (i.e. recording in SGSS) of positive microbiological samples for UK Biobank participants from 2015 onwards is 98%. However, prior to 2015 the reporting rates to SGSS varied markedly between local authorities.

### Detection of microbiological isolation in UKB participants [e.g.] is dominated by specific organisms, specimen types and regional laboratories

During 2015 (the year from which coverage for UKB participants in England can be considered almost complete), 21,361 individuals, corresponding to 4.79% of UK Biobank participants in England, had a positive microbiological culture recorded in SGSS. As expected from national data ^8,9,11,23^ *E. coli* (isolated from 1.93% of the cohort) and *S. aureus* (isolated from 0.67% of the cohort) were the commonest isolates, but major community and hospital associated pathogens were also represented, including *Enterococcus* spp., *Ps. aeruginosa*, various *Enterobacteriaceae* and *Streptococcus* spp. (Table 1).

**Table 1:**
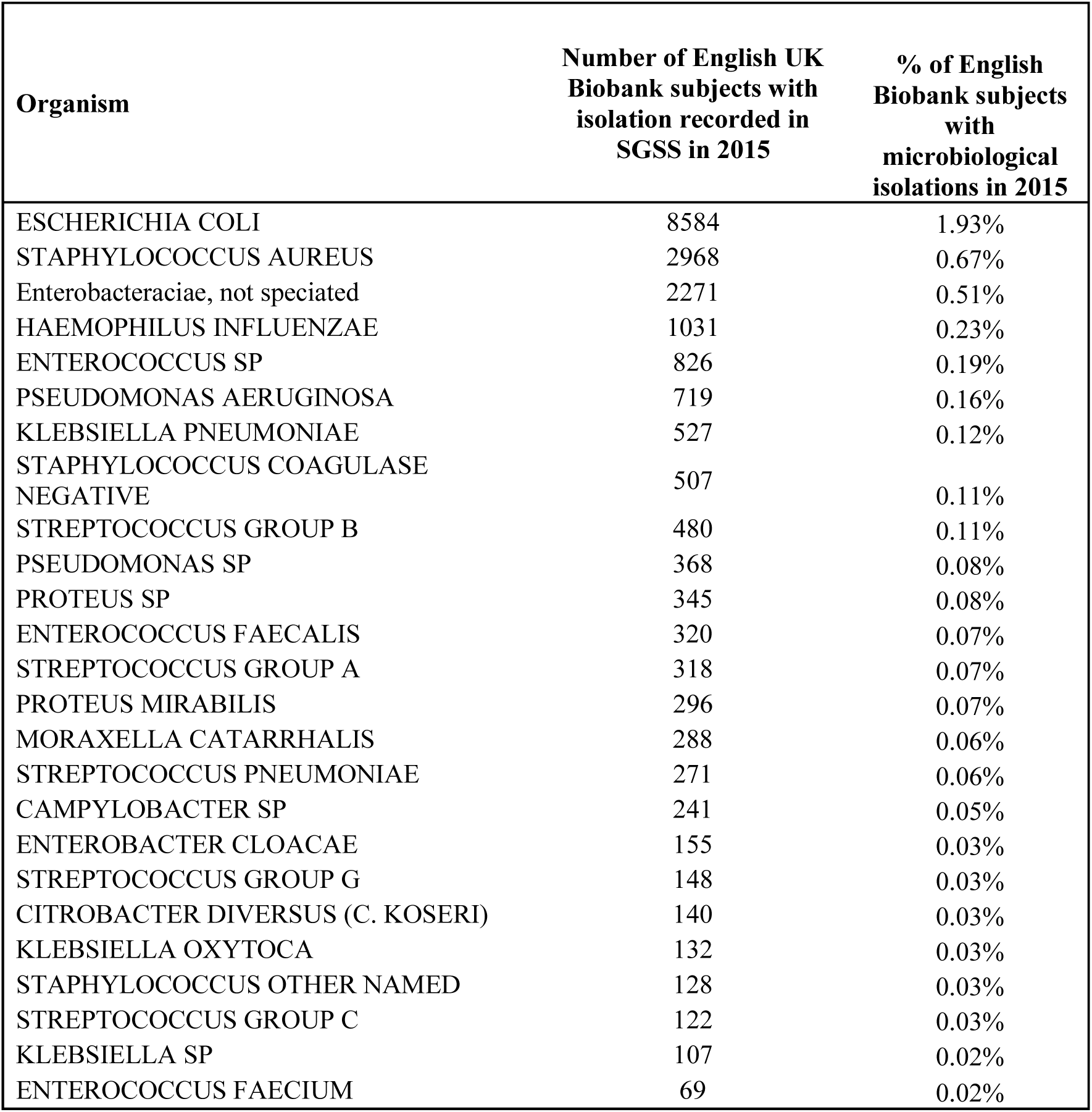
The number of UK Biobank participants with microbial isolations recorded in SGSS in 2015 for the 25 most common organisms isolated. Note that one individual’s cultures can yield more than one different microbe.

Isolates were derived from urinary, skin, sputum, blood, genital, and faecal samples, which are typical of current microbiological usage in NHS microbiology laboratories (Table 2). Urinary isolates were most common, with 12,468 individuals (2.80% of the English UK Biobank cohort) having a positive urinary isolate in 2015. By contrast, individuals with any positive blood cultures were relatively uncommon, with only 701 individuals with such isolates (0.16% of the cohort).

**Table 2:** The number of UK Biobank participants with samples recorded in SGSS in 2015, stratified by sample type from which the isolate was obtained.

| **Specimen type** | **Number of UK**<br>**Biobank subjects**<br>**with isolations**<br>**from Specimen**<br>**types recorded in**<br>**SGSS in 2015** | **% of English**<br>**Biobank subjects**<br>**with**<br>**microbiological**<br>**isolations in 2015** |
| --- | --- | --- |
| URINE/KIDNEY | 12468 | 2.80% |
| SKIN/WOUND | 2534 | 0.57% |
| SPUTUM | 1808 | 0.41% |
| BLOOD | 701 | 0.16% |
| SWAB | 649 | 0.15% |
| LOWER GENITAL TRACT | 533 | 0.12% |
| FAECES/LOWER GASTRO-INTESTINAL TRACT | 433 | 0.10% |
| NOSE | 413 | 0.09% |
| PUS SOURCE UNKNOWN | 242 | 0.05% |
| TISSUE | 181 | 0.04% |
| MIDDLE EAR/MASTOID | 190 | 0.04% |
| LOWER RESPIRATORY TRACT | 131 | 0.03% |
| UNKNOWN | 147 | 0.03% |
| FLUID – NOS | 122 | 0.03% |
| THROAT | 92 | 0.02% |
| EYE | 74 | 0.02% |
| INTRA-VASCULAR LINE | 44 | 0.01% |
| UPPER RESPIRATORY TRACT | 40 | 0.01% |

27% of the microbiological isolates from UK Biobank were recovered by five microbiology laboratories, 52% by twelve microbiology laboratories, and 70% by 21 laboratories (Table S1), which is relevant when considering the resource implications of obtaining microbial isolate specimens from UKB participants prospectively. Complete coverage by such a program would require participation of a large number of laboratories: 121 microbiology laboratories reported at least one specimen from a UK Biobank subject in 2015, 41 reported more than 100 specimens, 22 reported more than 300, while ten reported more than 600 specimens.

### Healthy participant effect

To qualify any possible “healthy participant” effect in infection outcomes, we analysed the last five quarters of the study period, as we considered the data most complete during this period (Figure 2). Table 3 shows isolation results for *E. coli, S. pneumoniae, S. aureus*, and *Campylobacter*, which are the most commonly isolated pathogens of urinary, respiratory, skin/wound, and bowel infection, respectively ^8,9,11,23^, as well as *Salmonella*, which is a rare isolate. Of these organisms, *E. coli* and *S. aureus* were by far the most common infections identified, with the majority isolated in primary care (Table 3).

**Table 3:** Numbers of individuals and (in brackets) rate per 1,000 persons years observed with isolation of various microorganisms. Numbers are presented stratified. Population refers to either the general population aged 40-69 during UK Biobank recruitment (Gen. Pop.), where the specimen was sent from (Facility = either Acute NHS Trusts (Acute) or from out of hospital settings (Community/General Practice (Comm/GP)).

| Population | FACILITY | Gender | *E. coli* | *S. pneumoniae* | *S. aureus* | *Campylobacter* | *Salmonella* |
| --- | --- | --- | --- | --- | --- | --- | --- |
| Most commonly infected site |  |  | Urine | Respiratory | Skin/wound | Bowel | Bowel |
| Gen. pop. | Acute | F | 201,267<br>(19.3) | 4,227 (0.4) | 107,600<br>(10.3) | 2,183 (0.2) | 584 (0.1) |
| **UKB** | **Acute** | **F** | **3,820**<br>**(14.0)** | **106 (0.4)** | **1653 (6.0)** | **51 (0.2)** | **17 (0.1)** |
| Gen. pop. | Acute | M | 66,402<br>(6.6) | 5,248 (0.5) | 154,101<br>(15.2) | 2,953 (0.3) | 618 (0.1) |
| **UKB** | **Acute** | **M** | **1,173 (5.1)** | **88 (0.5)** | **2224 (9.7)** | **57 (0.2)** | **12 (0.1)** |
| Gen. pop. | Comm/GP | F | 394,530<br>(37.9) | 5,244 (0.5) | 84,131 (8.1) | 4,733 (0.5) | 757 (0.1) |
| **UKB** | **Comm/GP** | **F** | **10,647**<br>**(38.9)** | **105 (0.4)** | **1339 (4.9)** | **162 (0.6)** | **22 (0.1)** |
| Gen. pop. | Comm/GP | M | 82,836<br>(8.2) | 4,880 (0.5) | 90,481 (8.9) | 5,817 (0.6) | 693 (0.1) |
| **UKB** | **Comm/GP** | **M** | **2,007 (8.8)** | **114 (0.5)** | **1518 (6.6)** | **205 (0.9)** | **24 (0.1)** |

Overall, rates of microbial isolation from UK Biobank participants are of similar order and pattern to those seen in the general population (Table 3,4). As expected, there is a higher rate of isolation of *E. coli* in women than in men (e.g. for samples sent from general practice settings, isolate rate was 38.9 in females vs. 8.8 in men per 1,000 person years observation), while *S. aureus* displays the opposite pattern (4.9 vs. 6.6 per 1,000 person years isolation, in women and men, respectively). For all *E. coli* isolated from primary care, overall isolation rates are similar in UKB participants and in the general population (Table 3, Table 4). In contrast, rates of *S. pneumoniae* and *S. aureus* isolation, and isolation of resistant *E. coli*, are slightly lower in UKB populations than in the general population, with incidence rate ratio estimates of 0.6 to 0.8 in different groups (Tables 3, 4). This effect is also seen in resistant *E. coli* isolates from primary care (Tables 3, 4).

**Table 4:** Incidence rate ratios (IRR) comparing isolation of *E. coli* (including resistant *E. coli* populations), *S. pneumoniae* and *S. aureus* from microorganisms recorded in SGSS April 2015 to August 2016, stratified by UK Biobank status. Microbiological isolations occur both in hospital (Facility = ‘Acute’) and out of hospital community settings (Facility = ‘Comm/GP’). Populations at risk of isolation differs between these populations: we used UK Biobank subjects as the denominator for the UK Biobank subjects, and the number of individuals 45-69 in mid 2015 as a denominator for non-Biobank cases.

|  |  | *E. coli* |  |  | *E. coli*, resistant to 3rd gen. cephalosporins |  |  | *E. coli*, resistant to ciprofloxacin |  |  | *S. pneumoniae* |  |  | *S. aureus* |  |  |
| --- | --- | --- | --- | --- | --- | --- | --- | --- | --- | --- | --- | --- | --- | --- | --- | --- |
| FACILITY | GENDER | 95% |  |  | 95% |  |  | 95% |  |  | 95% |  |  | 95% |  |  |
|  |  | IRR | CI |  | IRR | CI |  | IRR | CI |  | IRR | CI |  | IRR | CI |  |
| Acute | F | 0.72 | 0.70, | 0.75 | 0.94 | 0.91, | 0.97 | 0.84 | 0.56, | 0.6 | 0.96 | 0.78, | 1.16 | 0.59 | 0.56, | 0.61 |
| Acute | M | 0.78 | 0.74, | 0.83 | 0.73 | 0.70, | 0.77 | 0.84 | 0.80, | 0.87 | 0.74 | 0.59, | 0.91 | 0.64 | 0.61, | 0.67 |
| Comm/GP | F | 1.03 | 1.01, | 1.05 | 0.74 | 0.72, | 0.75 | 0.82 | 0.81, | 0.84 | 0.76 | 0.62, | 0.93 | 0.61 | 0.57, | 0.64 |
| Comm/GP | M | 1.07 | 1.02, | 1.12 | 0.78 | 0.75, | 0.82 | 0.83 | 0.79, | 0.86 | 1.03 | 0.85, | 1.24 | 0.74 | 0.70, | 0.78 |

Overall, these data support the idea that UK Biobank subjects are healthier than the general population, although the estimated healthy patient effect differs somewhat between organisms.

## DISCUSSION

We have demonstrated the feasibility of linking prospective cohort data (i.e. UK Biobank) with a national dataset containing information on microbial isolates in England (SGSS). The initiative was assisted by a clear ethical framework and common data model within both data sources, use of a proven pseudonymisation technology ^12^, and high personal identifier quality and utilisation (NHS numbers) in SGSS. The SGSS dataset has near complete (>98%) coverage of England from 2015 onwards and represents a single dataset containing microbial isolates from both primary and secondary care sources. It includes a data feed including all organisms on which antimicrobial susceptibility testing was performed, and the susceptibility results obtained. As microbiological standard operating procedures require that antimicrobial susceptibility testing be performed on clinically significant microbiological isolates ^24^, SGSS is likely to include a very high proportion of significant bacterial isolations in England.

Integration of microbiological data held by Public Health England with the UK Biobank study, the feasibility of which we have demonstrated here, has a number of potential advantages for public health and biomedical research. Although microbiological data obtained before 2015 is available in SGSS only in some areas of England, which reduces ascertainment by SGSS of isolation from UK Biobank participants in the period 2010-2015, at the time of writing, five full years (2015-2019) of nearly complete data can be linked, representing a large and powerful data source for epidemiological analysis.

However, before addressing these opportunities, we set out the limitations of the record linkage as implemented in this work.

Firstly, the UK Biobank cohort is generalizable to, but not necessarily representative of, the English population ^10^. Compatible with the healthy participant effect previously demonstrated in UK Biobank, microbial isolation rates were generally lower in UK Biobank participants than in the similarly aged population resident in the same area. For example, among women attending general practitioners, incidence rate ratios for *S. aureus, S. pneumoniae*, and *E. coli* resistant to ciprofloxacin in UK Biobank participants relative to the similarly aged population resident in the same area were 0.61 (95% CI 0.57, 0.64), 0.76 (95% CI 0.62,0.93), and 0.82 (0.81, 0.84) respectively. This likely reflects systematic differences in health care usage (such as use and selection of antibiotics) between UK Biobank and other subjects, but importantly, such biases can be quantified and used to interpret conclusions drawn UK Biobank – microbial data linkages.

Secondly, the validity of the data received by SGSS depends on the microbiological processing of the samples occurring in multiple labs. SGSS performs standardisation of nomenclature received from these diverse laboratories (term-mapping to a standard ontology). Across England there exists some heterogeneity between protocols and platform technologies used in different microbiological laboratories. This persists despite efforts to standardise practice, including the Standards for Microbiological Investigation which have been published and widely adopted in the UK ^24^, together with mandatory participation in external quality assurance schemes and periodic re-accreditation of laboratories. Therefore, some variation in results (for example, in antimicrobial susceptibility testing results, consequent on different methodologies being used) will exist across laboratories.

Thirdly, ascertainment of microbiological infection depends on access to medical care where relevant specimens are taken. Sampling policies may vary by medical practitioner ^25^. At present, SGSS only records the results of positive microbiological samples, so the rates of sample taking cannot be computed directly from SGSS data. In theory, comparison with data within UK Biobank derived from GP clinical systems (some of which now receive electronic copies of microbiology results, both positive and negative) might address this, as well as providing a route to cross-validate information derived from both GP systems and from SGSS. Monitoring the extent of microbial sampling with the UK Biobank cohort is an area for future work.

Finally, most infection is diagnosed based on symptoms and signs, without any contribution from microbiological investigation. Consequently, it is predominantly syndromic presentations which are coded and recorded in electronic record systems in primary and sometimes secondary care. Such diagnosis becomes much less accurate in older individuals ^1^. Compared with syndromic diagnosis, positive microbiological cultures from normally sterile sites have very high specificity^4,5^, with isolation recognised pathogens being indicative of severe infection in essentially all cases. By contrast, microbial isolation has a relatively low sensitivity for infection, particularly in mild disease and in respiratory infections, in which bacteraemia is detected in fewer than 20% of cases^3,26^. The specificity of microbial isolation from urine, which is much more common than isolation from blood cultures, is lower unless compatible symptoms are present^6,7^. It is high specificity which underlies the use of isolation of pathogens from blood cultures in infection surveillance programs^27,28^. Therefore, diagnoses of infection in the UK Biobank cohort using microbiological endpoints will allow specific identification of severe infections with a range of common organisms.

Nevertheless, the technical feasibility of monitoring microbiological isolations from over 450,000 individuals offers a number of important opportunities.

Firstly, because the UK Biobank contains genotyping on all of its participants, host genetics can be investigated as risk factors for microbial isolation for a wide range of organisms (Supplementary Table 1); it is likely that many host determinants of infection remain to be determined, e.g. ^20^.

Secondly, there are many pathogens, including *S. aureus, S. pneumoniae* and *N. meningitidis* to which the entire population is exposed^29^, but from which only a small proportion of subjects develop clinical disease. The basis of natural protection to these pathogens is poorly understood, and UK Biobank and similar cohort studies offer the opportunity to ‘learn from natural protection’ and thence identify pathogenic mechanisms, with concomitant downstream benefits such as the identification of novel biomarkers for diagnosis or vaccine targets for prevention.

The third opportunity concerns the monitoring of antimicrobial resistance using this platform. There is an expectation that large healthcare databases used can be used for continuous monitoring of population health^30^ ; monitoring the spread of antimicrobial resistance is one example^30^. UK Biobank now stores diagnosis and prescribing information from general practice consultations, through which 75% of the UK’s human antimicrobial exposure occurs^7^, as well as hospital inpatient data. This wealth of data on health outcomes, together with data from SGSS, will make it possible to analyse specific outcomes of interest (e.g. death, or hospital admission due to deterioration) associated with a given infection and taking into account co-morbidities, prior antimicrobial exposure (which selects for resistance), and the prior isolation of resistant microbes (which is a measure of resistance in the subject’s flora^29^), and of current antibiotic exposure. Such systems have the potential to monitor for *in vivo* antibiotic failure at a population level, a capability which does not currently exist at scale in the UK.

Additionally, new technologies enable identification of bacterial genetic elements and variants causally associated with virulence determinants, including novel antibiotic resistance elements ^31-34^. Such approaches have now been applied to a wide range of pathogens ^3536,37 383940^. As bacterial genome sequencing declines in cost, it has become possible to consider surveillance of pathogen populations being isolated from sites of infection at a genomic level; bacterial loci associated with virulence, spread, or antimicrobial resistance could all be identified by analyses co-modelling antimicrobial exposures and the other comorbidities, and for the identification of human-bacterial genetic interactions ^41^. The challenge is that specimen collection mechanisms would have to be established to do this. However, since 50% of the UK Biobank subjects’ samples are accrued in only 12 laboratories, using these laboratories as sentinel sites to gather microbial isolations as they are obtained, together with a program of centralised banking and sequencing, could be considered.

This approach, which is feasible given the record linkage process put in place here, would be globally unique. Inclusion of microbiological endpoints in the UK Biobank will allow this important resource to be used to address a range of important question related to infection in older adults.

## METHODS

### Collection of data from microbial laboratories by SGSS

All microbiological laboratories in England send data about identification of microbiological isolates to a central PHE database, the Second Generation Surveillance System. The SGSS dataset is updated on a daily basis via two data feeds. One contains mandatory reporting of a narrow range of pathogens of particular public health importance, including *Salmonella, Campylobacter* and other foodborne pathogens. A second data feed includes details of all microbial cultures on which antimicrobial susceptibility testing was performed. The antimicrobial susceptibility results transmitted to SGSS include all antimicrobial tests performed and their results, not just the clinically relevant subset reported by the laboratory to the clinician requesting the test. SGSS performs quality control checks and applies mappings between terms used by individual laboratories (including specimen types and microbiological species) to generate a standardised dataset.

We considered isolates received from individuals resident within English local authorities between 1 April 2010 and 30 June 2016, which we term the *study period*, unless otherwise stated. We made this restriction because coverage of Wales and Scotland by SGSS is not complete. We also restricted the comparative analysis of the general population to individuals of the same age range as that of UKB participants.

### Transfer and storage of data from UK Biobank

To establish which UK Biobank participants were also in SGSS, we used a system for encryption and storage of pseudonymised identifiers (OpenPseudonymiser^12^) to compare pseudo-anonymised (tokenised) NHS numbers present on SGSS records with the tokenised NHS numbers of UKB participants. This arrangement allows PHE to identify records from UK Biobank participants, but does not reveal to PHE the identity of the entire Biobank cohort. In exploratory analyses, additional identifiers (comprising date of birth, initial of forename and full surname, gender) were similarly tokenised to assess the feasibility of linkage of SGSS entries which lacked NHS numbers.

### Population estimates and computation of rates

Rates of recruitment and microbial isolation were estimated for each Local Authority (based on the participant’s address at recruitment), even when the address details provided to SGSS indicated they had moved. We used mid-year estimates of the population aged 40-69 resident in local authorities from which UKB recruited, stratified by gender, when comparing isolation rates in UKB subjects and in the general population. This data was obtained from the Office for National Statistics, UK.

### Healthy participant effect

Health outcomes and health-seeking behaviour differ between UK Biobank participants and the general population ^10^, with evidence of a “healthy participant” effect. To assess whether such effects also apply to infection outcomes, we compared isolation rates between UK Biobank participants and similarly aged individuals living in the English local authorities from which the UK Biobank recruited. We stratified isolation rates by gender, and by whether the specimen was received from hospital (secondary care) or from general practitioners (primary care).

### Ethical framework

PHE gathers data from NHS microbiology laboratories, storing it in the SGSS database. This data is fully identified, and anonymised extracts are generated prior to epidemiological analysis. This activity is permitted under Section 251 of the National Health Service Act 2006, which allows processing of named patient data without consent for defined purposes, including public health surveillance. Participants in the UK Biobank gave written, informed consent for UK Biobank to follow their health using linkage to electronic health-related records. Details of the ethical framework used by Biobank have been published ^22^.

## Data Availability

n/a

